# Venous Excess Ultrasound (VExUS) for Screening and Management of Acute Decompensated Heart Failure: A Prospective Observational Study

**DOI:** 10.1101/2025.09.17.25335674

**Authors:** AL Longino, GS Siegel, K Thayapran, DH Perez, A Manta, MD Kotzin, IS Douglas, JM Burke, E Gill, M Riscinti

## Abstract

**Background:** Bedside assessment of venous congestion is critical in the management of acute decompensated heart failure (ADHF. Conventional methods such as physical exam and laboratory tests have limited precision. Venous Excess Ultrasound (VExUS), a Doppler-based protocol assessing hepatic, portal, and renal venous flow, offers a promising, noninvasive method for evaluating systemic congestion.

**Objectives:** To evaluate the utility of VExUS in screening for cardiomyopathy, guiding diuresis, and predicting acute kidney injury (AKI) among emergency department (ED) patients admitted with suspected ADHF.

**Methods:** We conducted a prospective observational study at a public tertiary care center from July 2024 to April 2025, enrolling adults with suspected ADHF. Serial VExUS scans were performed from ED arrival through discharge. Primary outcomes included diagnosis of heart failure with reduced ejection fraction (HFrEF), AKI per KDIGO criteria, and change in serum creatinine following diuresis using multivariable regression and Receiver Operating Characteristic curves.

**Results:** Ninety-nine patients were included; 287 VExUS scans were analyzed. Admission VExUS grade predicted HFrEF (McFadden’s R2 = 0.3, p = 0.007) and correlated with both left ventricular ejection fraction (adjusted R2 = 0.3) and right ventricular dysfunction. VExUS grade decreased with net fluid loss (p < 0.001) and was associated with admission AKI (p = 0.01). Among patients diuresed with AKI, VExUS grade predicted creatinine trajectory (p = 0.01).

**Conclusions:** VExUS is a promising bedside tool for real-time assessment of venous congestion in ADHF. It may aid in diagnosis, guide diuretic therapy, and identify patients at risk for cardiorenal syndrome.

## Introduction

Heart Failure is a major contributor to morbidity, mortality, and healthcare costs around the world and in the United States.^1,2^ The 30-day mortality associated with ADHF is between 10 and 15%, on par with an advanced cancer diagnosis.^3^ While years of research spending and innovation have yielded improvements in long-term outcomes in these patients, serious outcomes have worsened in the United States, with a recent study highlighting recent increases in ADHF-associated mortality.^4^ Despite the severe consequences of ADHF, there are few evidence-based techniques for treatment of this common and deadly condition. The expansive 2022 American Heart Association (AHA) Heart Failure Guidelines provide few Class 1 recommendations for treatment of patients hospitalized with ADHF, the majority of which are based on limited data.^1^ Expert-informed guidelines recommend “…severity of congestion and adequacy of perfusion should be assessed…” and “Patients with HF with evidence of significant fluid overload should be promptly treated with intravenous loop diuretics to improve symptoms and reduce morbidity”, while allowing for initiation or titration of guideline directed medical therapy (GDMT).^1,5-8^ Cardiorenal syndrome (CRS) increasingly accompanies ADHF exacerbation, and is associated with higher rates of mortality and readmission,^9,10^ a negative prognostic indicator associated with a failure to achieve decongestion.^11,12^ Venous congestion is instrumental to the pathophysiology of CRS, and prompt treatment of venous congestion is key to improvement of clinical outcomes in patients with ADHF.^13^ Early detection of ADHF is critical for risk stratification and targeted management in the Emergency Department (ED).^14^ Thus, early, accurate, and ongoing assessment of venous congestion is key to the management of the undifferentiated ED patient, to inform diagnostic testing (e.g., with trans-thoracic echocardiography (TTE)) and management, balancing the need for decongestion with diuretics against the risk of over-diuresis leading to iatrogenic acute kidney injury (AKI).

Accurate bedside assessment of venous congestion is limited.^7,15-18^ Conventional physical exam maneuvers such as assessment of the jugular venous pulse suffer from low rates of inter-rater reliability and poor sensitivity and specificity for venous hypertension.^19^ Laboratory evaluations such as Brain Natriuretic Peptide (BNP) take time to result, and have known limitations in certain populations.^20^ Formal echocardiography and right heart catheterization, while more accurate than the physical exam, are time-consuming and costly procedures that are not easily conducted at the bedside, are not universally available, and cannot be repeated frequently to address questions of venous congestion. In daily practice, most providers rely on imprecise measures such as daily weights, changes in serum creatinine, or changes in the physical exam to guide diuretic management of patients hospitalized with ADHF. Even the currently accepted “gold standard” of invasive measurement of intracardiac pressures by right heart catheterization is a poor measure of venous flow, and is easily confounded by tricuspid regurgitation.^21^

For these reasons, there has been increasing interest in the use of bedside ultrasound to more accurately assess venous congestion to identify patients with ADHF, and to inform appropriate diuretic strategies.^19,22-24^ Initially promising ultrasound techniques such as measurement of the diameter inferior vena cava (IVC) have shown mixed results, leading to skepticism, and the need for more accurate, comprehensive bedside studies.^25-28^ One emerging ultrasound technique is Venous Excess Ultrasound (VExUS), a 4-step protocol integrating measurement of the IVC with Doppler assessment of flow through the hepatic, portal, and renal veins.^29^ Graded on a scale of 0 (no congestion) to 3 (severe congestion), the VExUS protocol has been shown to correlate closely with invasively-measured intracardiac pressures, AKI, and diuretic potential,^30,31^ to have a high degree of inter-rater reliability,^32^ and to track resolution of AKI in CRS.^33,34^ The relative simplicity and ease of interpretation of VExUS relative to formal echocardiography suggests that VExUS may be suitable as a means for ED or other bedside providers to rapidly assess a patient’s degree of venous congestion in order to make treatment decisions, or to screen for those who might benefit from further diagnostic studies such as transthoracic echocardiography (TTE) or right heart catheterization. Though there is a growing literature supporting the use of VExUS in ADHF,^35-40^ there has not yet been a thorough evaluation of the potential role of VExUS in early assessment and management of Emergency Department (ED) patients admitted to the hospital with suspected ADHF. To address this gap in the literature we conducted a prospective observational study in this key population evaluating the utility of VExUS to screen for clinically relevant cardiomyopathy, assess volume status, predict AKI, and guide fluid management.

We suspected that VExUS could be a useful tool to screen for ADHF and guide diuresis in a cohort of ED patients with suspected volume overload. Specifically, we hypothesized that VExUS grade would change with diuresis, providing a dynamic, real-time assessment of venous congestion; that VExUS could provide a rapid, point-of-care assessment to predict the likely utility of diuretics among patients with elevated serum creatinine; and that admission VExUS would predict which patients have decreased left or right ventricular (RV) function.

## Methods

Between 7/10/2024 and 4/1/2025 we conducted a prospective observational study following STROBE guidelines.^41^ We enrolled a non-sequential convenience sample of patients presenting to the ED of Denver Health Medical Center, a tertiary care center and academic-public safety-net hospital in Denver, Colorado. Inclusion criteria included age of 18 years or more, and a clinical suspicion of volume overload or ADHF on the part of the treating ED provider. Exclusion criteria included pregnancy, or the long-term inability to provide informed consent, as in cases of incarceration, chronic dementia, or chronic cognitive deficit. This observational study was approved by the

Colorado Multiple Institutional Review Board (COMIRB) and included waiver of informed consent for patients experiencing states of altered mental status such as intoxication, withdrawal, or delirium at the time of presentation.

On arrival to the ED, patients underwent standard initial assessment by ED providers. If there was clinical suspicion for ADHF, research staff were notified by the ED provider and the patients underwent serial VExUS scans on arrival to the ED prior to the initial dose of loop diuretic and on hospital days 1-3, and on the day of discharge.

### VExUS Scanning Protocol

the VExUS protocol was conducted as previously described, in accordance with recently published best-practice guidelines, with measurement of the IVC, and Doppler interrogation of the hepatic, portal, and renal vasculature.^29,39^ Patients were examined in a recumbent or semi-recumbent position using the curvilinear probe. All exams were conducted using the Mindray TE7 Max Ultrasound Machine (Shenzhen, China). All exams used electrocardiogram gating to increase accuracy of waveform interpretation.^32,39^ VExUS Sonographers included Emergency Medicine Ultrasound Fellows, as well as resident physicians, medical students, and research assistants that had undergone extensive training in the VExUS technique.

IVC Measurement: From the subxiphoid view the curvilinear probe was held in a craniocaudal orientation. The IVC was identified by entry into the right atrium, and the presence of the hepatic vein. The IVC diameter was measured perpendicular to the vessel, at the distal aspect of the hepatic vein insertion.

Hepatic Vein Doppler: Hepatic Vein flow was measured in any of its 3 branches.

Portal Vein Doppler: Portal venous flow was measured in the main, left, or right portal veins.

Renal Doppler: Renal flow was measured in the interlobar vessels, avoiding the renal hilum.

Interpretation: As previously described,^42,43^ the Doppler waveform of each vessel is determined to reflect a normal, moderate, or severe degree of congestion (appendix). If the IVC is less than 2 cm in diameter, the VExUS score is graded as 0, reflecting minimal congestion. If the IVC is greater than 2 cm and all waveforms are normal or mild, the VExUS grade is 1. If there is one severe waveform the VExUS grade is two, and if there are two or more severe waveforms, the VExUS grade is three, reflecting severe congestion. For purposes of quality control, interpreters gave each set of images an assessment of image quality, evaluated on a 1–5 scale recommended by the American College of Emergency Physicians, and images with a quality score of less than 3 were discarded.^44^ For image interpretation, deidentified images were reviewed independently by two members of the research team with experience interpreting VExUS exams. When disagreement occurred, experts jointly reviewed deidentified images and adjudicated a final score. Ultrasound images were recorded and scored using the Butterfly Network Blueprint Quality Assurance Software (Burlington, MA).

### VExUS Training

All scanners watched a one-hour VExUS training video reviewing the technique in detail. Scanners then underwent a minimum of two two-hour in-person training sessions to further refine their skills before starting to collect data. Training sessions continued throughout the study period.

### VExUS Grading

All doppler images were blinded and then reviewed by two expert VExUS interpreters to generate a consensus determination of image quality and VExUS grade. In the event of disagreement, joint review was conducted. If needed, a third expert VExUS interpreter could be asked to make a final ruling.

### Data Extraction

Manual data extraction was performed from the Epic electronic medical record (Verona, Wisconsin). Data collected included demographic information, presence and resolution of AKI per KDIGO criteria,^45^ inpatient and 30-day mortality, need for supplemental oxygen, echocardiographic findings, lab results, and daily fluid balance, with particular focus on capturing all creatinine measurements and diuretic doses administered. 2-D Echocardiograms were performed when clinically indicated at the discretion of the treating provider. Echocardiograms were performed by certified cardiac sonographers and interpreted by board-certified cardiologists. Echocardiographic data were extracted from the reading cardiologist’s report, including Left Ventricular Ejection Fraction (LVEF), Right Ventricle (RV) function, RV dilation, E/e’ ratio, and presence of valvular disease.

For statistical purposes, RV function and dilation were coded on a numeric scale of 0-4 corresponding to values of “none”, “mild”, “moderate” or “severe”. Data extraction was performed by two medical staff trained in chart review. All elements of the Charlson Comorbidity Index (CCI) were collected, and the CCI was independently calculated for each patient.^46^ Patient data were stored in a REDCap database (Nashville, TN). The data dictionary and codebook used for data extraction are available as supplemental materials.

All study procedures were approved by the local Institutional Review Board (COMIRB # 22-2024).

### Statistical Methods

Descriptive characteristics on patient demographics and outcomes of interest were tabulated and are displayed in table 1. Data are presented as Mean (SD) unless indicated. For purposes of logistic and linear regression, VExUS grade was coded as an ordinal variable, from 0-3. The covariates of age, sex, and CCI were selected a-priori based on clinical significance and existing literature).^46^ Correlation analyses of VExUS grade and NT-ProBNP with LVEF tertile were conducted using linear regression controlling for age, sex, and CCI. Association of VExUS grade and NT-ProBNP with presence of LVEF <40% was analyzed using logistic regression controlling for age, sex, and CCI. We then used ROC analysis to assess area under the curve (AUC) for VExUS grade and NT-ProBNP for diagnosis of Heart Failure with reduced ejection fraction (HFrEF), as well as a combined model. Youden indexing was used to determine the ideal cut point and sensitivity and specificity of VExUS grade for detection of HFrEF. The association between RV function and RV dilation was assessed using linear regression controlling for age, sex, and CCI, with RV function and dilation coded as continuous variables as described above. Associations between VExUS grade and hospital day and change in VExUS grade and change in net fluid balance were assessed using linear regression. Association between admission AKI and VExUS grade was conducted using logistic regression controlling for age, sex, and CCI. To assess the relationship between change in creatinine in response to diuresis, we identified patients that met admission criteria, presented with an elevated creatinine, and underwent diuresis on arrival to the ED. Among those patients we used linear regression to evaluate the association between initial VExUS grade and change in the next creatinine value measured following diuresis. Patients with missing or uninterpretable admission VExUS scans were excluded from the analysis without imputation for missing clinical or imaging data.

## Results

Between 7/10/2024 and 4/1/2025 we screened 129 patients. Of these, 13 were excluded due to not meeting inclusion criteria or declining to provide informed consent. Of the remaining 116 patients, 17 were excluded from the final analysis due to poor image quality. 287 scans were included in the final analysis. Initial disagreement on scan interpretation requiring joint review occurred in 24 (8%) of included images. In all cases a consensus was reached without involvement of a third interpreter.

### Demographics

Of the 99 patients enrolled, the average age was 63 (53, 71), and 59 (60%) were male. Fifty-nine (60%) required supplemental oxygen on admission. Mean Admission NT-ProBNP was 5,241 ng/L (2,482, 11,149); mean admission high-sensitivity troponin was 38 ng/L (14, 82) ; mean admission lactate was 2.15 mmol/L (1.35, 3.15). 75 (85%) had an AKI on arrival to the ED. Forty-nine (50%) had a history of myocardial infarction and 86 (88%) had a known history of congestive heart failure.

### Cardiac Echocardiographic Features

Valvular disease was common in the cohort, with 64 (65%) patients having some degree of tricuspid regurgitation and 21 (21%) having some degree of mitral regurgitation. The mean left ventricular ejection fraction (LVEF) was 40% (25, 56), and the mean E/e’ ratio was 14 (11, 19).

### Cardiomyopathy Screening

Among patients that underwent TTE during the index admission (n = 60), VExUS grade was significantly associated with a diagnosis of HFrEF (McFadden’s R^2^ = 0.3, p = 0.007), as well as with same-admission left ventricular ejection fraction tertile (adjusted R^2^= 0.3, p = 0.001) after controlling for age, sex, and Charlson Comorbidity Index. Of note, in the same patients, admission N-terminal Pro Brain Natriuretic Peptide (NT-ProBNP) was not associated with HFrEF diagnosis (p = 0.1) but was associated with Ejection fraction tertile (Adjusted R^2^= 0.15, p = 0.04) (Figure 2).

**Figure 1.**
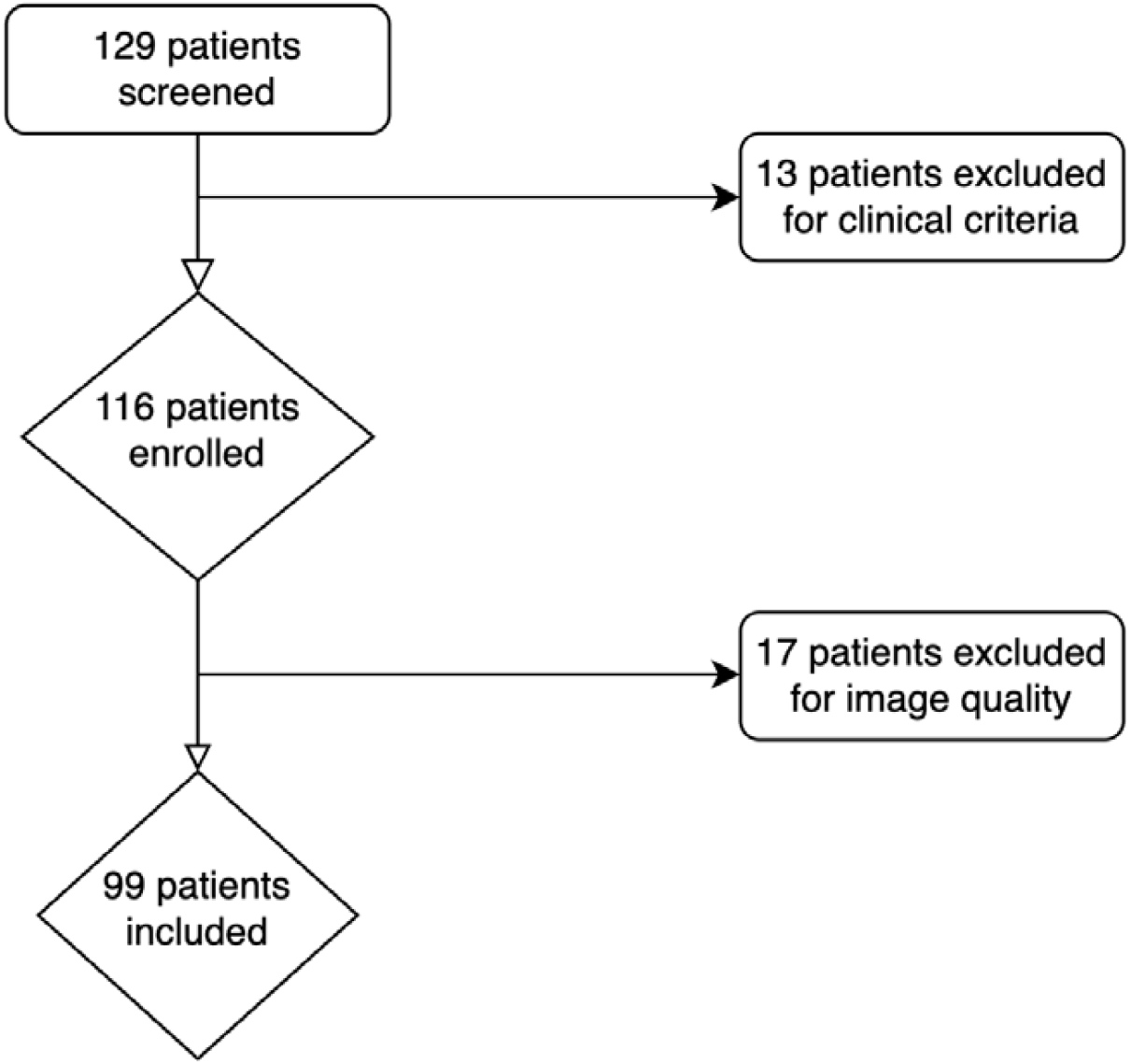
Patient Enrollment Diagram

**Figure 2.**
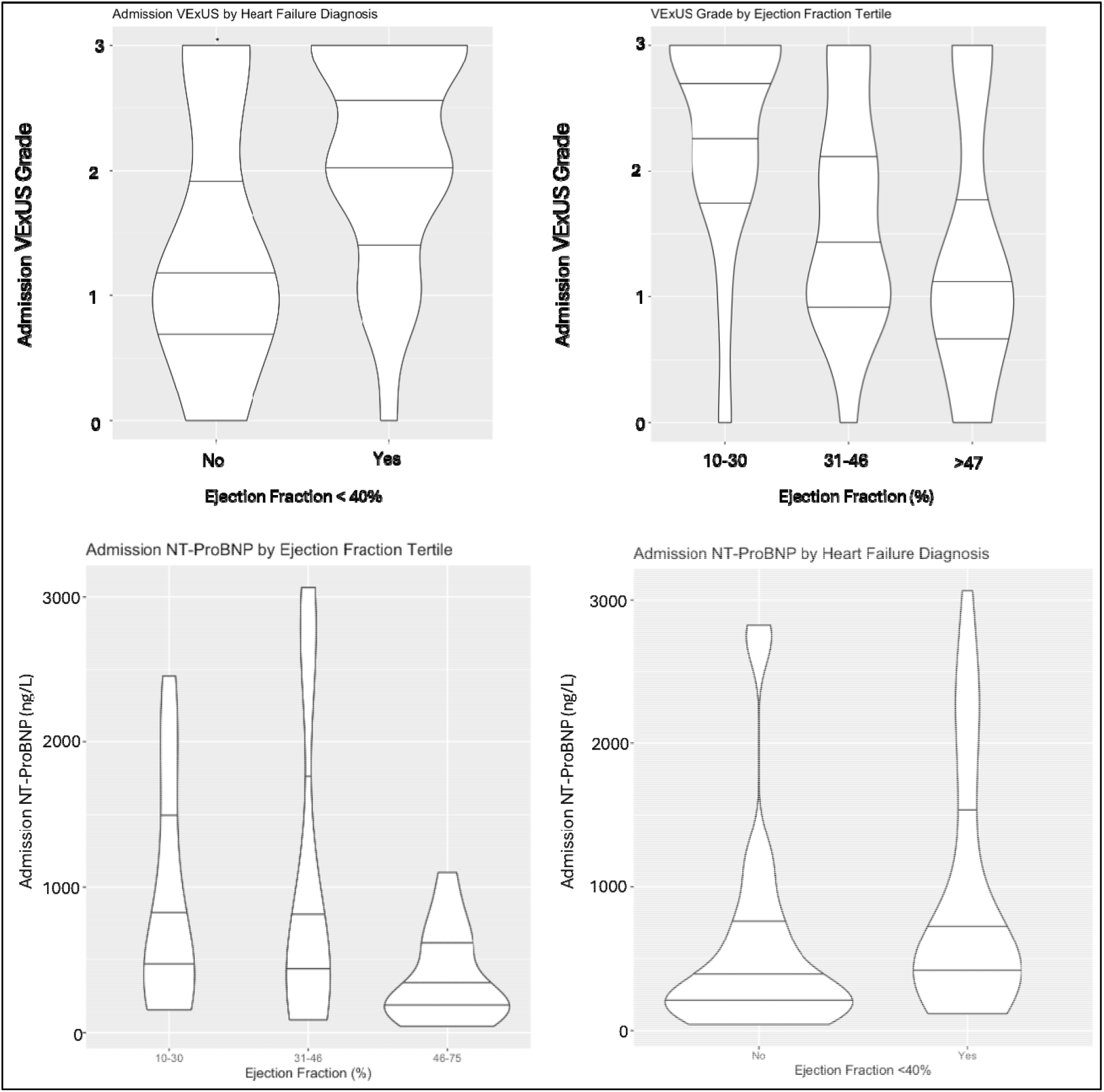
Admission VExUS Grade is associated with same-admission diagnosis of Heart Failure with Reduced Ejection Fraction (McFadden’s R^2^ = 0.3, p = 0.007) and Ejection Fraction tertile (adjusted R^2^ = 0.3, p = 0.001) after controlling for age, sex, and Charlson Comorbidity Index. Admission NT-ProBNP was not significantly associated with Heart Failure with Reduced Ejection diagnosis (p = 0.1), and was significantly associated with Ejection Fraction Tertile (Adjusted R^2^= 0.15, p = 0.04).

In a Receiver Operating Characteristic analysis comparing VExUS and NT-ProBNP for the detection of patients with a same-admission TTE with an ejection fraction <40%, controlling for age, sex and Charlson Comorbidity Index, VExUS demonstrated an elevated area under the curve (AUC) (0.76, 95% CI 0.65 – 0.87), relative to NT-ProBNP (AUC 0.68, 95% CI 0.56 – 0.81). The optimal cut point for VExUS was a grade of 1, found to have a sensitivity of 0.76 and a specificity of 0.68 (Figure 3). Of note, a model including both NT-ProBNP and VExUS grade had an AUC of 0.8 (95% CI 0.71 – 0.91).

**Figure 3.**
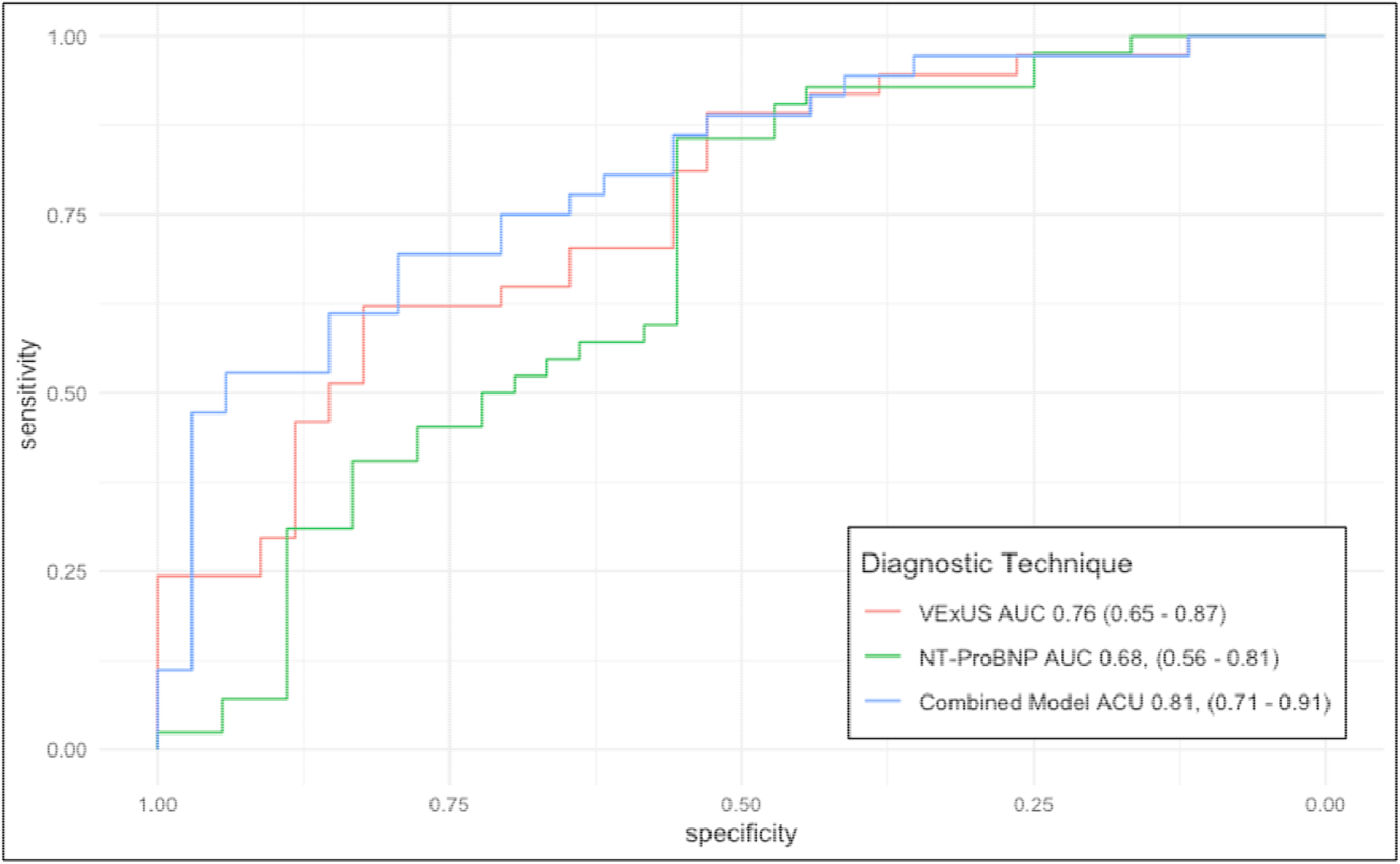
Receiver Operating Characteristic Curve for VExUS and NT-ProBNP for the detection of patients with a same-admission TTE with an ejection fraction <40%, controlling for age, sex and Charlson Comorbidity Index. VExUS demonstrated an elevated AUC (0.76, 95% CI 0.65 – 0.87), relative to NT-ProBNP (AUC 0.68 95% CI 0.56 – 0.81). A model including these covariates, NT-ProBNP and VExUS grade had an AUC of 0.8 (95% CI 0.71 – 0.91).

Comparison of VExUS Grade and NT-ProBNP for Prediction of Heart Failure with Reduced Ejection Fraction VExUS grade was associated with both right ventricular dilation (Adjusted R^2^= 0.26, p = 0.0005) and decreased right ventricular function (Adjusted R^2^= 0.24, p = 0.001) after adjusting for age, sex, and Charlson Comorbidity Index (Figure 4).

**Figure 4.**
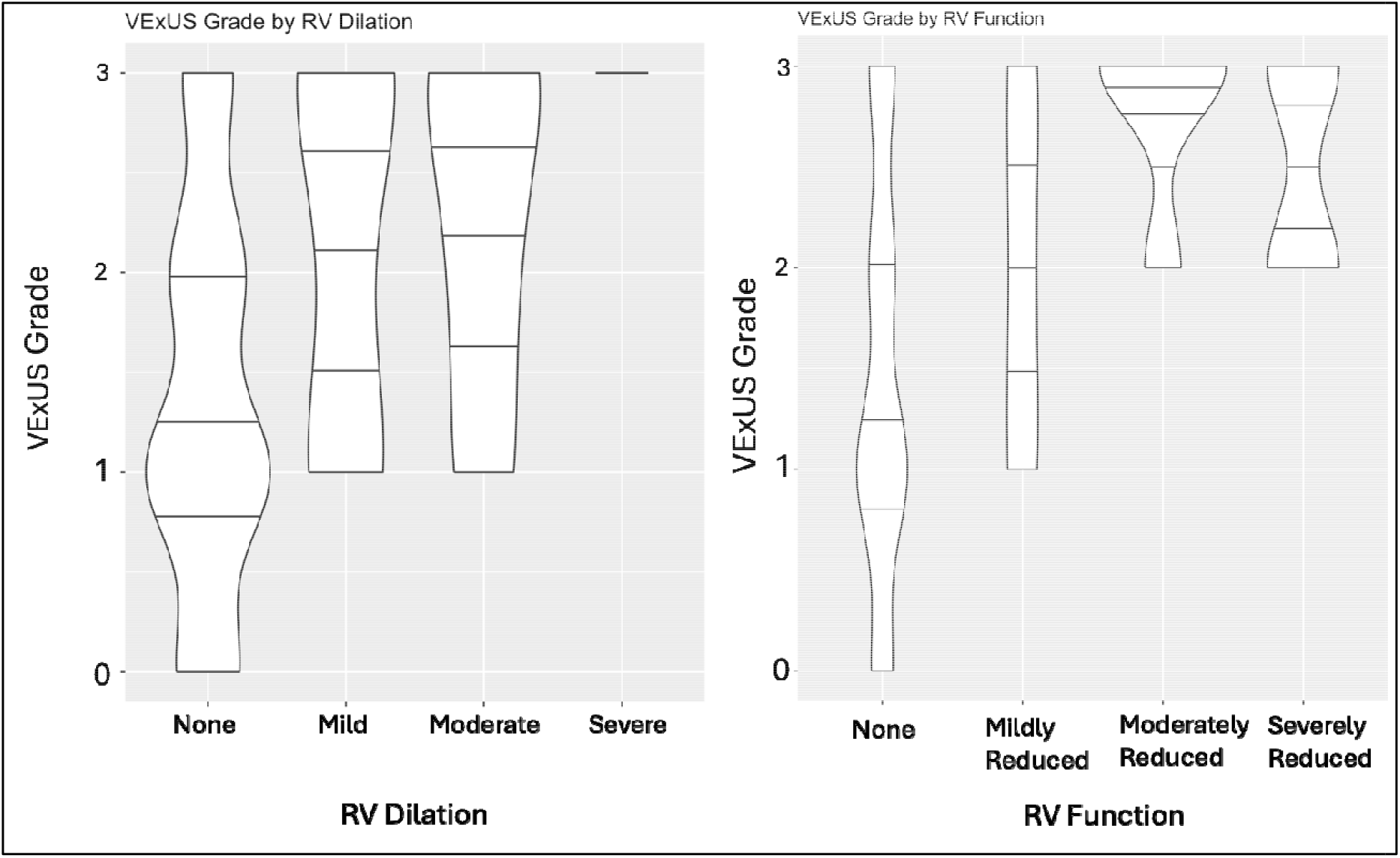
VExUS grade was associated with both right ventricular dilation (Adjusted R2= 0.26, p = 0.005) and decreased right ventricular function (Adjusted R2= 0.24, p = 0.001) after adjusting for age, sex, and Charlson Comorbidity Index

**Figure 5.**
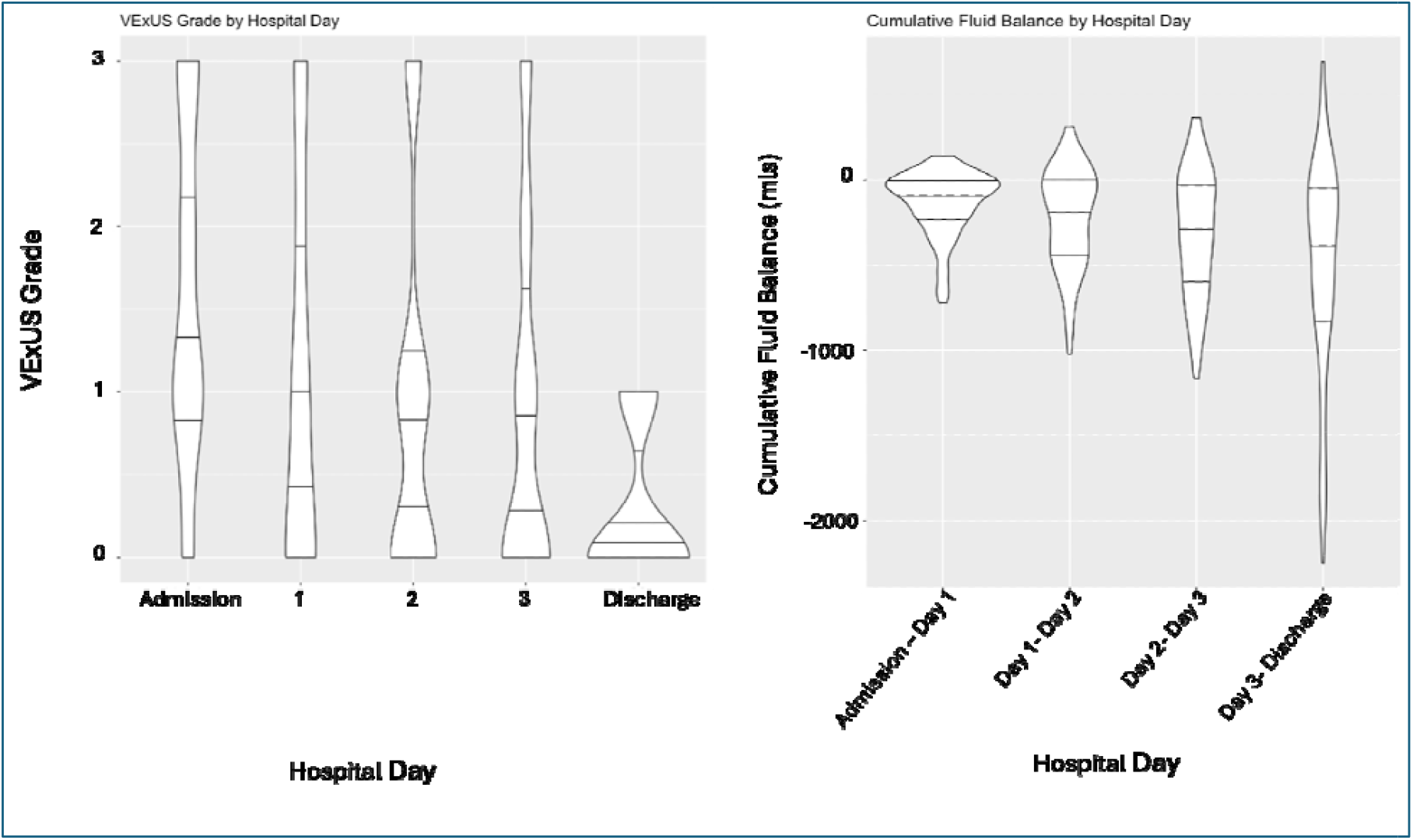
VExUS grade showed a statistically significant linear decrease over time (Adjusted R^2^ = 0.01, p <0.001), that was significantly associated with change in fluid balance (adjusted R^2^ = 0.02, p = 0.04).

**Figure 6.**
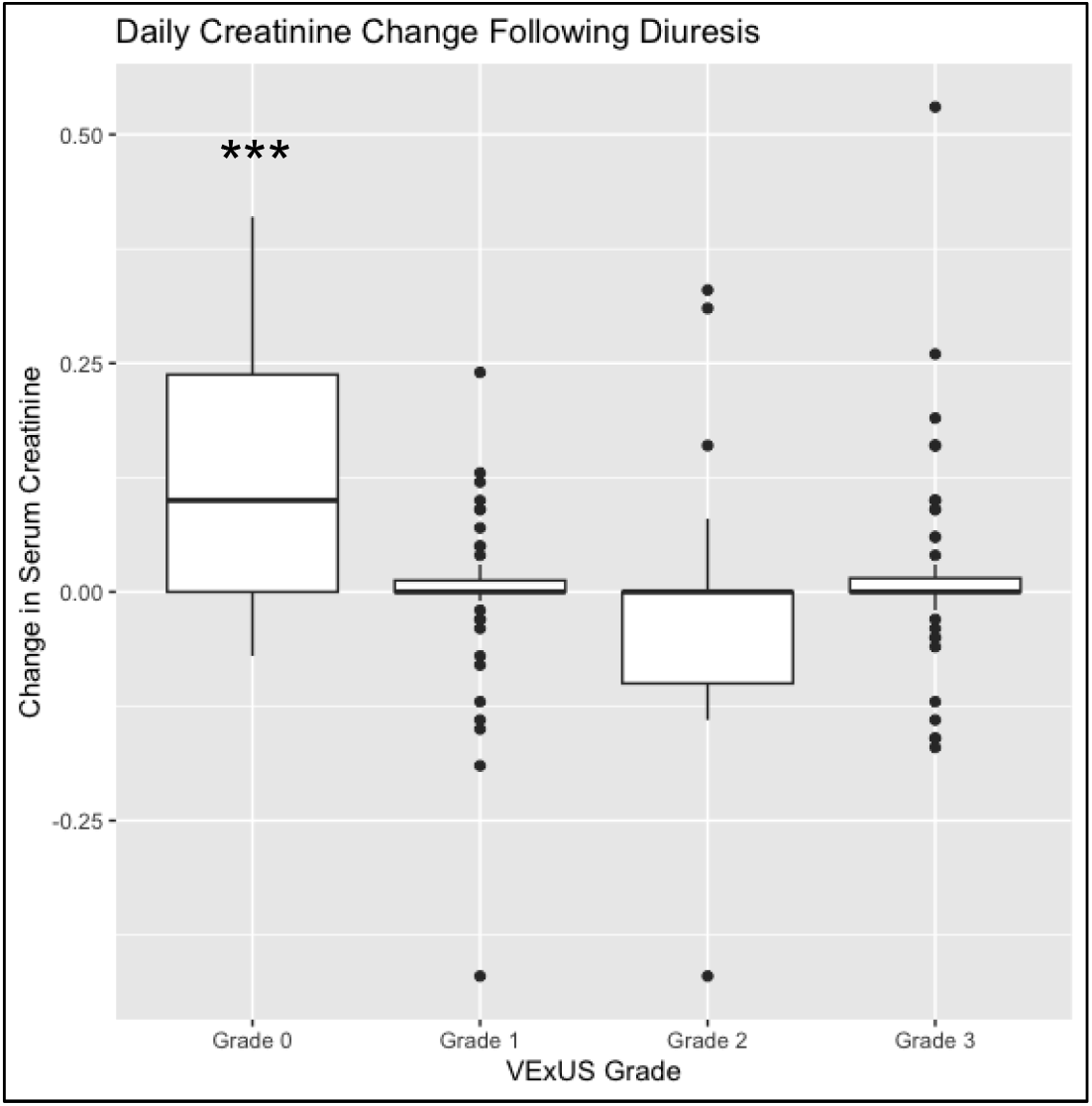
Among patients presenting with AKI that underwent diuresis, there was a significant indirect association between elevated VExUS grade and subsequent change in serum creatinine (Adjusted R^2^ = 0.07, p = 0.01).

There was no significant association between admission VExUS grade and either Heart Failure with Preserved Ejection Fraction diagnosis or E/e’ ratio on same-admission TTE.

### Fluid Balance

VExUS Grade demonstrated a statistically significant linear decrease over time as patients underwent diuresis (Adjusted R^2^ = 0.01, p = <0.001). This change was significantly associated with change in fluid balance, as measured by trained nursing staff tracking fluid input and output (Adjusted R^2^ = 0.02, p = 0.005).

### Acute Kidney Injury and Response to Diuresis

After adjusting for age, sex, and CCI, admission VExUS grade was associated with the presence of acute kidney injury on admission, per KDIGO criteria(McFadden’s R^2^ = 0.17, p = 0.01).^45^ Additionally, among patients presenting with AKI that underwent diuresis on admission (n = 67), a VExUS Grade of 0 was associated with a subsequent increase in serum creatinine (Adjusted R^2^ = 0.07, p = 0.01).

## Discussion/Conclusion

Our findings suggest that Doppler assessment using VExUS may be highly informative for patients hospitalized with ADHF by rapidly providing diagnostic information at multiple timepoints during a patient’s hospital stay. Sequential changes in VExUS correlating with fluid balance indicate a sensitive functional read-out for therapeutic response monitoring to diuresis in ADHF. Considering the 2022 AHA Heart Failure guideline’s Class 1 recommendation that “severity and congestion and adequacy of perfusion should be assessed to guide triage and initial therapy”,^1^ a rapid, point-of-care ultrasound exam that can be feasibly performed by a triaging clinician may have a valuable role to play in identifying patients at risk for complications of ADHF.

### Cardiomyopathy Screening

VExUS grade demonstrated a similar AUC to NT-ProBNP when identifying which patients would demonstrate a reduced ejection fraction on same-admission TTE (figure 3) suggesting that VExUS may be a valuable bedside screening tool, providing diagnostic information to providers in real time, before laboratory results become available, or in whom laboratory results might be confounded. Furthermore, the fact that a model including both VExUS and NT-ProBNP had an elevated AUC relative to either VExUS or NT-ProBNP alone may suggest that VExUS could be a valuable adjunct to current laboratory screening tests for cardiac dysfunction in the ED. Similarly, the association of VExUS with RV dilation and dysfunction may prove to be an important signal to providers to consider right-sided pathology, or prompt a provider to order a formal TTE in a patient that might otherwise be misdiagnosed. Interestingly, while other studies have shown associations with RV dysfunction and dilation, elevated risk for in-hospital mortality and readmission among patients with known HFrEF,^40,47^ we believe this to be the first study to suggest that VExUS may be an efficient screening tool for clinically significant left sided cardiomyopathy in the ED population.

### Acute Kidney Injury and Diuretic Management

The current study’s findings of an association between elevated VExUS grade and admission AKI in a hypervolemic cohort reconfirms findings of prior VExUS studies showing associations between elevated VExUS grade and cardiorenal AKI among patients with heart failure.^29-31,33,48^ Bhardwaj et al specifically demonstrated that in addition to predicting the presence of AKI in CRS, VExUS can be used to track and predict resolution of cardiorenal AKI.^33^ Perhaps most interestingly, we demonstrate that VExUS grade may be able to predict renal tolerance of diuretic therapy. Given the highly-morbid and complex nature of CRS, coupled with the fact that not all hypervolemic patients with elevated creatinine have CRS,^10^ VExUS may be able to help determine which patients presenting with AKI are most likely to benefit from diuretic administration. These results support the findings of a recent review of VExUS for CRS monitoring and treatment supporting its use.^34^ Aggressively identifying and treating CRS in the ADHF population is critical, given the increased mortality associated with renal dysfunction in this vulnerable population.^49^ Importantly, identifying patients with right of left-sided cardiac dysfunction that might benefit from diuresis before lab results are available or a formal TTE can be completed has the potential to decrease “door to diuretic time”, allowing for earlier initiation of loop diuretics, a strategy associated with a 23% reduction in 30-day mortality in a recent metanalysis (OR 0.77; 95% CI 0.64-0.93).^50^ The finding that daily change in VExUS grade had a significant association with daily change in fluid balance aligns with multiple recent studies of VExUS in ADHF patients,^22,33,36,47^ suggests that VExUS may be a useful bedside tool for monitoring volume status over the course of a patient’s used predict a patient’s likely response to diuresis, assisting bedside providers throughout the hospital stay with decision-making around frequency and quantity of diuretic dosing in order to avoid iatrogenic kidney injury from over-diuresis, reducing patients’ length of stay.

There are several strengths of our study including a well-characterized, clinically relevant, generalizable cohort of emergency department patients with fluid overload, a focus on patient-centered outcomes, expert interpretation of results, and description of a novel and cost-effective bedside technique for CHF screening and management.

There are several important limitations, including the relatively small sample size of the cohort, the single-center enrollment and non-sequential patient enrollment. The academic center in which the study was carried out has a robust ultrasound training program with multiple providers familiar with VExUS technique, which may limit the external validity of the reported findings. Furthermore, VExUS is a novel technique, and as such there are limited studies validating its findings. However, there have been several studies describing both its correlation with invasively-measured hemodynamic variables,^30,31^ and its relatively high degree of inter-rater reliability.^32^. Another consideration is that echocardiograms were obtained based on clinical need rather than per research protocol, raising the possibility that patients with lower clinical concern for structural cardiac disease may not have been captured in the cardiomyopathy screening analysis.

In summary, VExUS represents a potentially widely available, non-invasive and relatively cost-effective technique in patients hospitalized with ADHF, providing insight into cardiomyopathy screening, diuretic tolerance, and dynamic fluid status monitoring. The findings require validation and confirmation in related populations. Further efforts to standardize and automate ultrasound protocols including image acquisition and interpretation, through enrollment of larger numbers of patients, and harmonize research aims and protocols. Use of VExUS to predict iatrogenic AKI from overdiuresis, the use of VExUS to inform decision making around timing of patient discharge, as well as the outpatient use of Doppler-based examinations for diuretic titration.

### Clinical Perspectives

The current study suggests that the novel VExUS technique may be play a significant role in the daily ED and inpatient management of ADHF, either as a means of screening patients for cardiomyopathy, or assisting with management decisions around diuresis. However, much remains unknown about the technique, including how easily it can be taught, and what utility it can demonstrate in randomized study designs. Future studies should focus on broader implementation of VExUS-driven diuresis protocols.

## Data Availability

Data will be made available on request.

