## Supplementary figures and images for "Venous Excess Ultrasound (VExUS) for Screening and Management of Acute Decompensated Heart Failure: A Prospective Observational Study"

### Appendix

Appendix A:

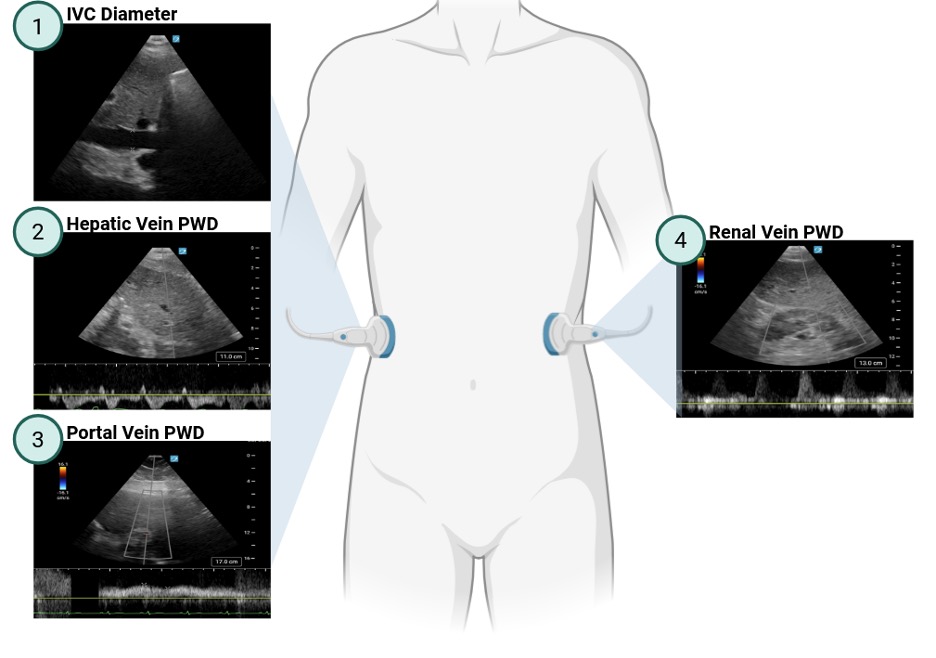

Appendix B:

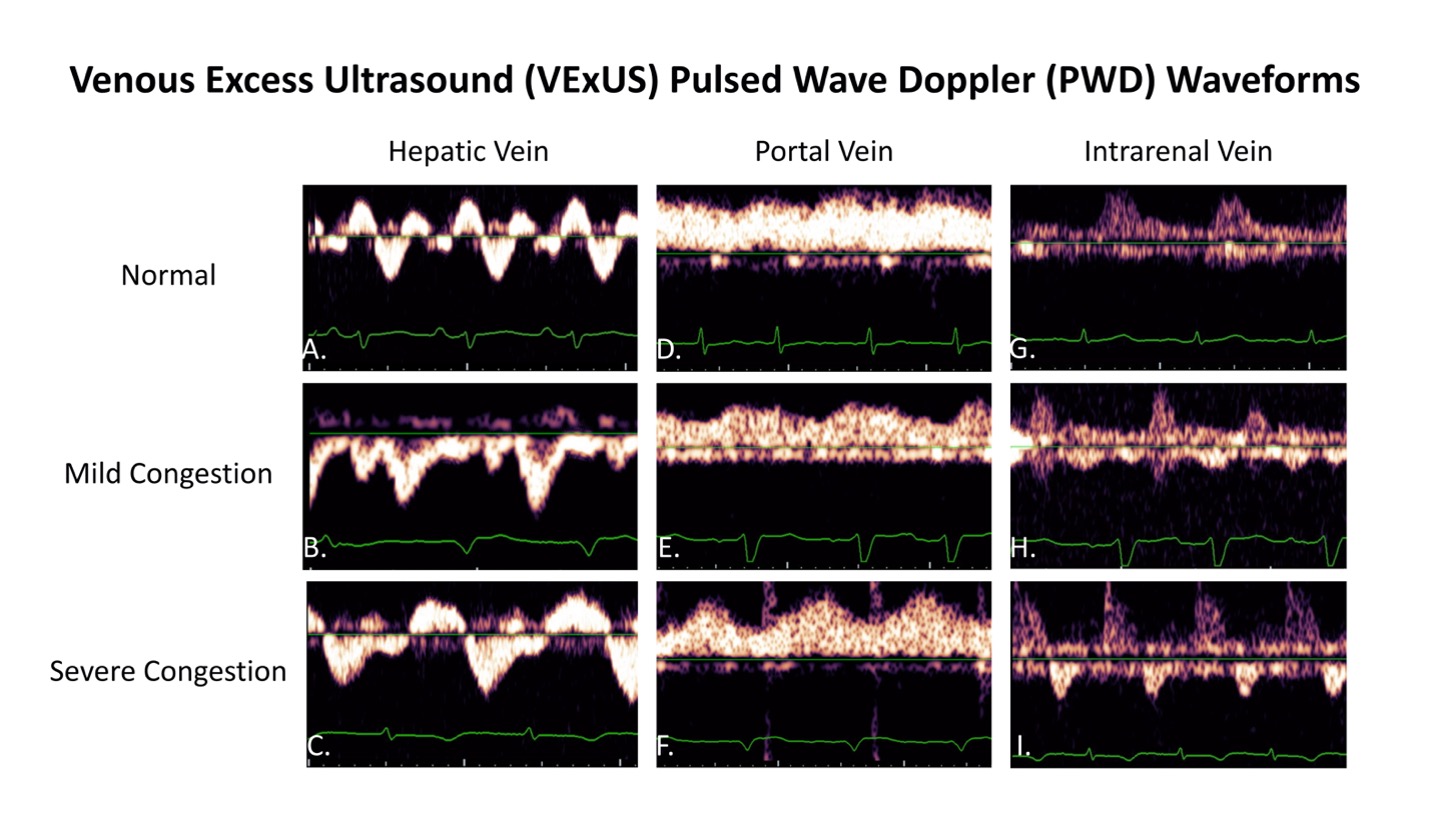
