## Supplementary material for "Venous Excess Ultrasound (VExUS) for Screening and Management of Acute Decompensated Heart Failure: A Prospective Observational Study": Table 1

| Characteristic | N = 99*^1^* |
| --- | --- |
| Age | 63 (53, 71) |
| Sex |  |
| Male | 59 (60%) |
| Female | 39 (40%) |
| Race |  |
| Asian | 1 (1.0%) |
| Black or African American | 20 (20%) |
| Native Hawaiian | 1 (1.0%) |
| Other | 10 (10%) |
| White | 67 (68%) |
| Ethnicity |  |
| Hispanic | 37 (38%) |
| Non-Hispanic | 61 (62%) |
| Admission MAP | 106 (86, 119) |
| Requiring Supplemental Oxygen | 59 (60%) |
| Admission Glasgow Coma Score |  |
| 11 | 1 (1.0%) |
| 13 | 3 (3.0%) |
| 14 | 4 (4.0%) |
| 15 | 90 (91%) |
| 16 | 1 (1.0%) |
| Admission NT-ProBNP (ng/L) | 5,241 (2,482, 11,149) |
| Admission HS-Troponin (ng/L) | 38 (14, 82) |
| Admission Lactate (mmol/L) | 2.15 (1.35, 3.15) |
| Admission Platelet Count (× 109/L) | 228 (171, 274) |
| *1*  Median (Q1, Q3); n (%) |  |

| Characteristic | N = 99*^1^* |
| --- | --- |
| Admission Bilirubin (mg/dL) | 0.80 (0.35, 1.30) |
| Admission AKI |  |
| Absent | 75 (85%) |
| Present | 13 (15%) |
| History of Myocardial Infarction | 49 (50%) |
| History of Congestive Heart Failure | 86 (88%) |
| History of Peripheral Vascular Disease | 15 (15%) |
| History of Stroke or TIA | 22 (22%) |
| History of Dementia | 13 (13%) |
| History of COPD | 50 (51%) |
| History of Liver Disease |  |
| No | 86 (89%) |
| Mild | 8 (8.2%) |
| Moderate | 2 (2.1%) |
| Severe | 1 (1.0%) |
| History of Diabetes |  |
| No | 54 (55%) |
| Controlled | 20 (20%) |
| End-Organ Damage | 24 (24%) |
| Left Ventricular Ejection Fraction | 40 (25, 56) |
| RV Function |  |
| Normal | 45 (60%) |
| Mildly Reduced | 11 (15%) |
| Moderately Reduced | 15 (20%) |
| Severely Reduced | 2 (2.7%) |
| Abnormal, Unspecified | 2 (2.7%) |
| *1*  Median (Q1, Q3); n (%) |  |

| Characteristic | N = 99*^1^* |
| --- | --- |
| RV Dilation |  |
| None | 38 (52%) |
| Mild | 16 (22%) |
| Moderate | 12 (16%) |
| Severe | 6 (8.2%) |
| Abnormal, Unspecified | 1 (1.4%) |
| Mitral Valve Regurgitation | 65 (66%) |
| Trace MR |  |
| Absent | 73 (74%) |
| Present | 26 (26%) |
| Mild MR |  |
| Absent | 78 (79%) |
| Present | 21 (21%) |
| Moderate MR |  |
| Absent | 78 (79%) |
| Present | 21 (21%) |
| Severe MR |  |
| Absent | 96 (97%) |
| Present | 3 (3.0%) |
| Tricuspid Regurgitation |  |
| Absent | 35 (35%) |
| Present | 64 (65%) |
| Tricuspid Regurgitation Severity |  |
| Trace | 35 (55%) |
| Mild | 16 (25%) |
| Moderate | 11 (17%) |
| *1*  Median (Q1, Q3); n (%) |  |

| Characteristic | N = 99*^1^* |
| --- | --- |
| Severe | 2 (3.1%) |
| Aortic Regurgitation |  |
| Absent | 72 (73%) |
| Present | 27 (27%) |
| Degree of Aortic Regurgitation |  |
| Trace | 19 (70%) |
| Mild | 5 (19%) |
| Moderate | 2 (7.4%) |
| Severe | 1 (3.7%) |
| Average E/e' Ratio | 14 (11, 19) |
| Heart Failure With Reduced Ejection Fraction | 41 (54%) |
| Heart Failure with Preserved Ejection Fraction | 29 (47%) |
| Heart Failure with Moderately Reduced Ejection Fraction | 29 (47%) |
| *1*  Median (Q1, Q3); n (%) |  |
